# Dosing of convalescent plasma and hyperimmune anti-SARS-CoV-2 immunoglobulins: a phase I/II dose finding study

**DOI:** 10.1101/2023.03.07.23286893

**Authors:** Sammy Huygens, Tim Preijers, Francis H. Swaneveld, Ilona Kleine Budde, Corine H. GeurtsvanKessel, Birgit C.P. Koch, Bart J.A. Rijnders

## Abstract

**Background:** During the COVID-19 pandemic, trials on convalescent plasma (ConvP) were performed without preceding dose-finding studies. This study aimed to assess potential protective dosing regimens by constructing a population pharmacokinetic (popPK) model describing neutralizing antibody (Nab) titers following the administration of ConvP or hyperimmune globulins(COVIg).

**Methods:** Immunocompromised patients, testing negative for anti-SARS-CoV-2 spike antibodies despite vaccination received a range of anti-SARS-CoV-2 antibodies in the form of COVIg or ConvP infusion. The popPK analysis was performed using NONMEM v7.4. Monte Carlo simulations were performed to assess potential COVIg and ConvP dosing regimens for prevention of COVID-19.

**Results:** 44 patients were enrolled, and data from 42 were used for constructing the popPK model. A two-compartment elimination model with mixed residual error best described the Nab-titers after administration. Inter individual variation was associated to CL (44.3%), V1 (27.3%), and V2 (29.2%). Lean body weight and type of treatment (ConvP/COVIg) were associated with V1 and V2, respectively. Median elimination half-life was 20 days (interquartile-range: 17–25 days). Simulations demonstrated that even monthly infusions of 600ml of the ConvP or COVIg used in this trial would not achieve potentially protective serum antibody levels for >90% of the time. However, as a result of hybrid immunity and/or repeated vaccination plasma donors with extremely high Nab-titers are now readily available, and a >90% target attainment should be possible.

**Conclusion:** The results of this study may inform future intervention studies on the prophylactic and therapeutic use of antiviral antibodies in the form of ConvP or COVIg.

## Introduction

Since the start of the pandemic, COVID-19 has taken millions of lives.(1) Effective vaccines can now prevent severe COVID-19 disease, hospitalization and mortality.(2-4) Unfortunately, a heterogeneous group of patients (e.g. those with solid organ transplant, hematological malignancies, or with anti-CD20 therapy) still have a poor or completely absent humoral immune response after primary vaccination as well as boosters.(5) They continue to be at risk for a prolonged and/or severe COVID-19 disease.(6)

By mid-2021, several monoclonal virus neutralizing antibodies (mAbs) had become available as a treatment in parts of the world and can also be used as pre-or post-exposure prophylaxis.(7, 8) MAbs target one specific epitope in the spike protein. Unfortunately, subsequent SARS-CoV-2 variants accumulated mutations in these epitopes which resulted in loss of activity against these variants.(9, 10)

In contrast to mAbs, polyclonal antibodies (pAbs) may be less susceptible to changes in the spike protein.(11, 12) Both convalescent plasma (ConvP) and hyperimmune globulins (COVIg) are forms of pAbs. ConvP is plasma from donors who have recovered from or were vaccinated against SARS-CoV-2.(13) COVIg is an intravenous immunoglobulin product produced from pooled plasma from more than 1000 donors and included ConvP donations.(14) The main advantage of ConvP is that it can be collected very early on in a pandemic, but its antiviral activity varies between each donor. In contrast, it takes several months to produce a first batch of COVIg, but it is more polyclonal than ConvP, the antibody content is constant in each vial of a batch, and ABO blood group matching is not required.

An unprecedented number of trials on the efficacy of ConvP and a few as well on COVIg as a treatment for COVID-19 were completed during the first 24 months into the pandemic.(15, 16) The results of these trials have been contradictory. As with mAbs, most evidence in favor of ConvP has been generated in patients very early after symptom onset and in the context of immunodeficiency.(17, 18) More importantly, several animal studies and a recent meta-analysis on outpatient ConvP therapy showed that a high enough SARS-CoV-2 spike antibody titer is essential to observe a therapeutic benefit.(16, 19, 20) MAbs as well as pAbs may also be used to prevent SARS-CoV-2-virus infections in immunocompromised patients who lack an endogenous antibody response after vaccination. However, dosing regimens of ConvP or COVIg that result in a potentially protective neutralizing antibody (Nab) titer for a minimum duration of e.g. 28 days are unknown because proper dose-finding studies with ConvP and COVIg remain unavailable.

This study aimed to establish a population pharmacokinetic (PK) model that is able to predict Nab-titers obtained after infusion of SARS-CoV-2 antibodies using ConvP or COVIg and can be applied to assess potential protective dosing regimens.

## Methods

### Study design

A single-center, open label, phase I/II prospective non-randomized trial (Trial NL9379) was conducted at the Erasmus Medical Center (Rotterdam, the Netherlands). The protocol was approved by the Dutch competent authority (CCMO) and the institutional review board (METC) at Erasmus MC. Written informed consent was obtained from all patients.

Based on the availability of ConvP and later of COVIg, the inclusion of a total of 104 patients was planned across groups of different doses and products. In figure S1, the study design is presented in detail. Treatment allocation (ConvP/COVIg) was open-label. These study arms were further divided into different volumes and concentrations. The batches with high Nab-titers were tested first. Patients in the ConvP group were allocated to a predefined volume and Nab-titer based on ABO compatibility. Patients included in the COVIg arm could participate a second time in a ConvP arm of the study at the time they had become SARS-CoV-2 spike antibody negative.

### Study products

ConvP was provided by the Dutch bloodbank (Sanquin Blood Supply). Donors met the standard plasma donor criteria, had a history of symptomatic COVID-19, and had recovered for at least 14 days. COVIg was manufactured by Prothya Biosolutions and provided by the Dutch Ministry of Health, Welfare and Sport. These particular batches were derived from pooled plasma from at least 1000 donors, including ConvP donations. Both products were produced while the ancestral variant (Wuhan-1) type was dominant in the Netherlands and, therefore, the Nab-titer against this strain was measured. The methods of the Nab-titer measurement are described in the supplementary methods (S2). Antibody treatment was administered intravenously. Since ConvP was collected before anti-SARS-CoV-2 vaccines had become available, plasma with very high Nab-titers was rare and the majority of the ConvP from these non-vaccinated donors had a Nab-titer ranging between 270 and 500 IUmL^-1^. In this study, ConvP with a Nab-titer of 500 and 910 IUmL^-1^ was used and is referred to as intermediate-titer and high-titer ConvP, respectively. By pooling regular plasma with ConvP, Prothya was able to produce two batches of COVIg with an increased Nab-titer of 270 and 910 IUmL^-1^. These products will be referred to as low-titer and high-titer COVIg, respectively. The Nab-titer, given in IUmL^-1^, is a unit of antibody neutralization of the ancestral SARS-COV-2 variant, as described by Nguyen et al. It facilitates the comparison of Nab-titers between a broad range of in-house virus neutralization tests.(21)

Because IgG titers correlated well with neutralization assays of the ancestral virus, titers of IgG antibodies against the spike protein measured with the LIAISON^®^ SARS-CoV-2 TrimericS IgG assay (DiaSorin) were used.(21). The LIAISON^®^ SARS-CoV-2 TrimericS IgG assay comprised a chemiluminescence immunoassay (CLIA) determining the anti-trimeric spike protein-specific IgG antibodies. Results of the CLIA are reported in binding affinity units/mL (BAUmL^-1^), which is the preferred unit for binding capacity by the WHO.(22-24) CLIA was performed on 11 of the 13 administered ConvP units from which median binding capacities were 3230 BAUmL^-1^ and 3070 BAUmL^-1^ for the intermediate and high Nab-titer, respectively. CLIA was performed ten times on the high Nab-titer COVIg batch (910 IUmL^-1^), from which a median of 3985 BAUmL^-1^ was obtained. For a rough estimate of the Nab-titer in IUmL^-1^, the result of the LIAISON^®^ SARS-CoV-2 TrimericS IgG assay can be divided by 4.

### Patient selection

Patients were at least 18 years old and did not have anti-SARS-CoV-2 antibodies at baseline. First, patients who had received B-cell depleting therapy were included but after the start of the vaccination campaign, all patients lacking anti-SARS-CoV-2 antibodies at least two weeks after full vaccination (two mRNA vaccines, two adenovirus vector vaccines (ChAdOx1-S), or one adenovirus vector vaccine (Ad26.COV2.S)) could participate in the study as well. Patients were screened with a point-of-care antibody test (Roche SARS-CoV-2 Rapid Antibody Test^®^). Negative test results were verified by the DiaSorin CLIA test and were deemed negative if Nab-titers were <33.8 BAUmL^-1^ according to the manufacturer’s instructions.(23) Patients had no symptoms of SARS-CoV-2 infection and tested negative with a qPCR test at the time of screening for the study.

### Clinical and biochemical monitoring

SARS-CoV-2 spike antibody measurement was performed using CLIA at baseline and, subsequently, after 24 and 48 hours and after 1, 2, 4, 6, 8, 12, 18, and 24 weeks or until the Nab-titer had become negative (<33.8 BAUmL^-1^) again. Blood sampling was also halted if the patient received another anti-SARS-CoV-2 vaccination during follow-up or had a breakthrough infection.

### Primary endpoints

#### Population pharmacokinetic analysis

To perform a population PK analysis, the measured Nab-titers versus time curves from ConvP and COVIg were described using non-linear mixed-effect modeling with NONMEM v7.4 (ICON Development Solutions, Ellicott City, MD, USA), which was guided using PsN v4.9.0. Pirana v2.9.9 was used for model management and R v4.2.1 (R Core Team, 2020) with Xpose v4.5.3 were used for graphical model diagnostics.(25-27) For obtaining the model parameters, first-order conditional estimation (FOCE) was applied with epsilon-eta interaction.

#### Model development

Model development commenced with evaluating the most parsimonious compartment model to describe the Nab-titer versus time data with initial parameter values obtained from the literature.(28, 29) For constructing the statistical model, the residual unexplained variability (RUV) was evaluated using an additional, proportional, or mixed (additive and proportional) error model. Moreover, inter-individual variability (IIV) was evaluated for each parameter separately using a log-normal distribution. Inter-occasion variability (IOV) was not estimated, as only data from one dosing event was collected. Model building was conducted using a stepwise approach, as the addition of parameters to the model was evaluated one by one.

#### Covariate analysis

In the covariate analysis, the patient and treatment characteristics were used to explain the obtained IIV for the model PK parameters. Selection of the covariate relationships was based on biological and clinical plausibility. For evaluating the dichotomous covariate relationships, the following model was applied:

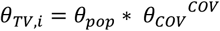

in which *θ*_*TV,i*_ is the typical value for the individual patient *i*, θ_pop_ is the population PK parameter value, *θ*_*cov*_ the parameter describing the covariate effect and *COV* is the covariate value being 1 if present and 0 otherwise. For the continuous covariate relationships, the following relationships (linear, power, and exponential) were applied:

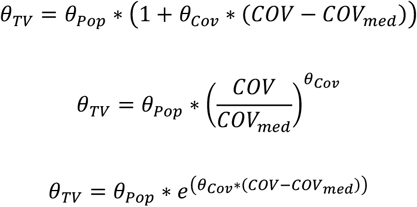

in which *COV*_*med*_ is the median for the covariate value. Before applying a covariate model, the plausibility of that relationship was first evaluated using graphical exploration. The covariate model was constructed using a standard forward inclusion (*p*=0.05, df=1) and backward elimination (*p*=0.01, df=1) procedure.

#### Model evaluation

The ability of the model to predict the Nab-titer measurements was described using an objective function value (OFV). As the OFV is χ_2_ distributed, the difference between the OFVs (dOFV) from two hierarchical models was used for model selection and dOFV values of 3.84 and 6.64 indicated a significant difference of *p*<0.05 and *p*<0.01 for one degree of freedom, respectively.

Model evaluation and selection were also based on graphical exploration using goodness-of-fit (GOF) plots, and prediction-corrected visual predictive checks (pcVPCs). Furthermore, the robustness of the parameter estimation from the final model was evaluated by non-parametric bootstrap analysis with 2000 replications.

#### Dosing regimen simulation

To verify which dosing regimen would result in predefined Nab-titer targets, Monte Carlo simulations of different dosing regimens were conducted using the final model. For the covariate relationships of the final model, values were taken randomly from the study cohort. Dosing regimens were rounded to the nearest practical volume.

#### Secondary endpoints

To evaluate the protective effect of ConvP and COVIg, patients were instructed to undergo PCR testing when they would become symptomatic in order to detect potential breakthrough SARS-CoV-2 infections. If possible, the viral strain was sequenced for patients admitted to the hospital. For investigating the safety of ConvP and COVIg, serious adverse events (SAE) were assessed.

## Results

### Patient population and follow-up

Patients were screened for eligibility between April 2021 and April 2022. The study was terminated prematurely due to the emergence of the Omicron variant (B.1.1.529) which became the dominant variant in early 2022 in the Netherlands.(30) Indeed, as the ConvP and COVIg available for the study had a much lower Nab-titer against this B1.1.529 variant, we did not expect any further potential benefit from study participation for the individual patient.

In total, 60 patients were screened and 44 were enrolled in the study (Figure 1). Patients were allocated to the intermediate and high Nab-titer groups first. One patient in the COVIg group was excluded from further analysis since the confirmatory anti-SARS-CoV-2 antibody test turned out positive at baseline before dose administration. In addition, one patient from the ConvP group was excluded since this patient accidently received plasma without SARS-CoV-2 neutralizing antibodies. Demographics and baseline characteristics were similar for both treatment groups (Table 1 and Table S3).

**Figure 1.**
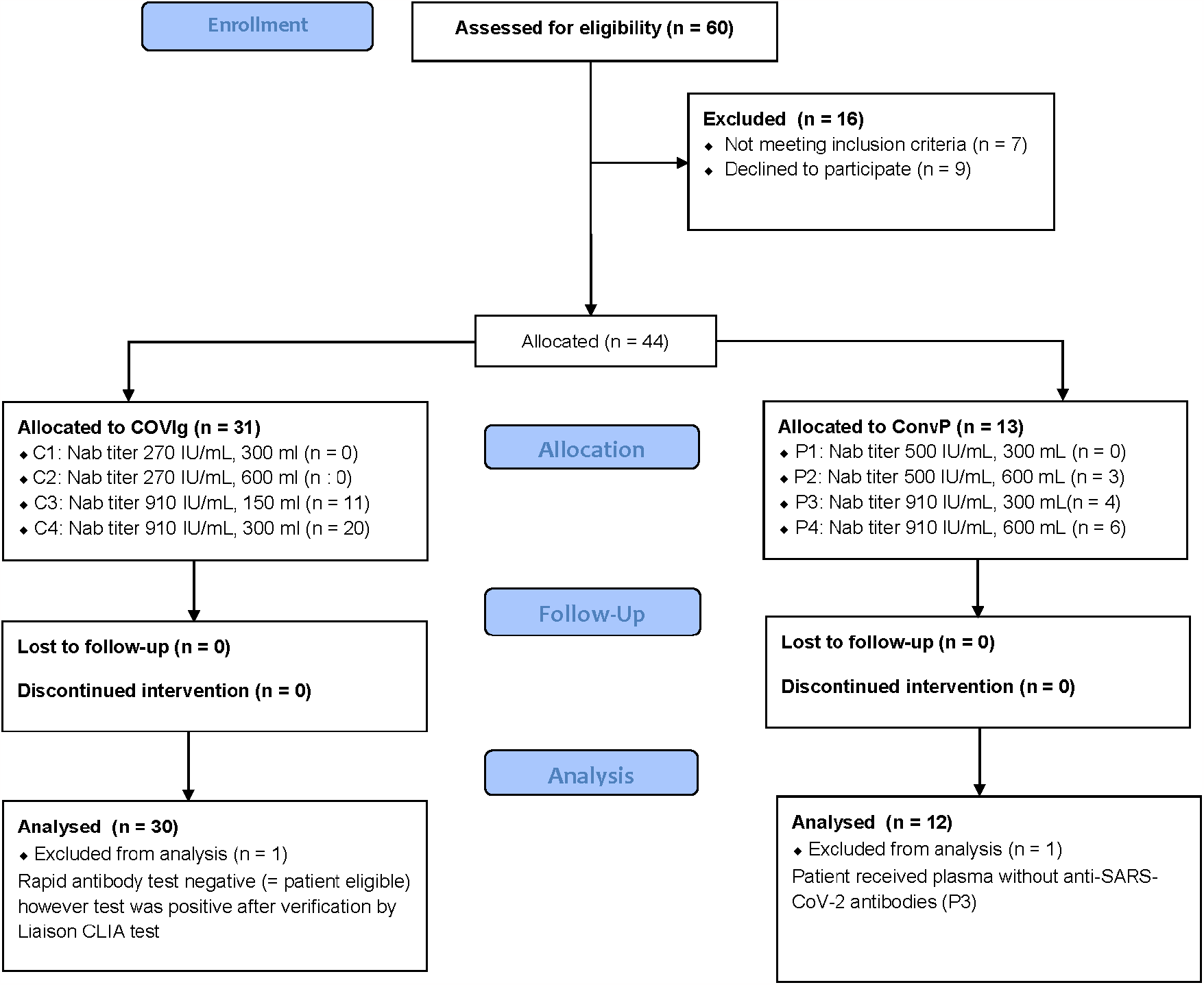
CONSORT flow diagram: patient enrollment and allocation of treatment. For each of the eight treatment cohorts (P1 to P4 and C1 to C4), the neutralizing antibody titers and the administered volume are depicted. N depicts the number of patients assigned to the corresponding treatment cohort. Abbreviations: CLIA = chemiluminescent immuno-assay; ConvP = convalescent plasma; COVIg = hyperimmune globulins containing anti-SARS-CoV-2 antibodies.

**Table 1.** Demographics and baseline characteristics of the total patient population and treatment groups.

| Baseline characteristics | Total<br>n = 42 | ConvP<br>n = 12 | COVlg<br>n = 30 |
| --- | --- | --- | --- |
| Age (yr) – median (IQR) | 61 (51 – 64) | 60 (46 – 64) | 62 (51 – 64) |
| Male gender – no. (%) | 22 (52.4) | 5 (41.7) | 17 (56.7) |
| Ethnicity – no. (%) |  |  |  |
| - Caucasian | 37 (88.1) | 10 (83.3) | 27 (90) |
| - African | 2 (4.8) | 1 (8.3) | 1 (3.3) |
| - Asian | 2 (4.8) | 1 (8.3) | 1 (3.3) |
| BMI (kgm<sup>2</sup>) – median (IQR) | 30 | 26 (23 – 32) | 28 (23 – 31) |
| Body weight (kg) – median (IQR) | 82 | 81 (68 – 88) | 84 (73 – 97) |
| Body length (cm) – median (IQR) | 178 (167 – 183) | 178 (164 – 183) | 178 (168 – 184) |
| Cause for immunocompromised state – no. (%) |  |  |  |
| - Rituximab | 26 (61.9) | 8 (66.7) | 18 (60.0) |
| - SOTx | 7 (16.7) | 1 (8.3) | 6 (20.0) |
| - Ocrelizumab | 4 (9.5) | 2 (16.7) | 2 (6.7) |
| - HSCT | 2 (4.8) | - | 2 (6.7) |
| - CVID | 1 (2.4) | 1 (8.3) | - |
| - Other reasons | 2 (4.8) | - | 2 (6.7) |
| Vaccination status – no. (%) |  |  |  |
| - Full vaccination<sup>a</sup> | 38 (90.5) | 10 (83.3) | 28 (93.3) |
| - Third vaccination<sup>b</sup> | 8 (18.1) | 4 (33.3) | 4 (13.3) |
| Laboratory findings - median (IQR) |  |  |  |
| - Hematocrite (%) | 42.5 (38 – 45) | 41.5 (37.3 – 45) | 43 (40 – 45) |
| - Leukocytes (μL<sup>-1</sup>) | 5550 (4275 – 8450) | 4450 (3850 – 6800) | 6300 (4450 – 8650) |
| - Lymphocytes (μL<sup>-1</sup>) | 1045 (768 – 1253) | 1000 (788 – 1225) | 1085 (768 – 1313) |
| - B-lymphocytes (μL<sup>-1</sup>) | 0 (0 – 40) | 0 (0 – 0) | 0 (0 – 75) |
| - T-lymphocytes (μL<sup>-1</sup>) | 790 (595 – 1225) | 730 (660 – 1170) | 810 (560 – 1230) |
| - CD4+ cells (μL<sup>-1</sup>) | 530 (355 – 715) | 530 (330 – 710) | 510 (358 – 723) |
| - CD8+ cells (μL<sup>-1</sup>) | 250 (155 – 485) | 220 (160 – 470) | 300 (148 – 563) |
| - NK cells (μL<sup>-1</sup>) | 190 (95 – 275) | 260 (80 – 290) | 175 (98 – 263) |

Abbreviations: ConvP = convalescent plasma; COVlg = hyperimmune anti-SARS-Cov-2 globulins; CVID = common variable immune deficiency; HSCT = hematopoietic stem cell transplant; IQR = interquartile range; NK = natural killer; SOTx = solid organ transplant
<sup>a</sup> Two mRNA vaccines, two vaccines by AstraZeneca or one Johnson & Johnson.
<sup>b</sup> Second vaccination (mRNA) in case of vaccination with Johnson & Johnson.
<sup>c</sup> Mann-Whitney U test was performed for continuous variables. Chi-square test was performed for categorical variables.
<sup>d</sup> Fisher's exact test was performed since low amount of observations in at least one cell.

In total, 86% of the patients (36/42) could be followed until SARS-COV-2 spike antibodies had become negative again. Antibody measurement was halted in three patients because of a breakthrough infection, and in one patient due to an antibody response after an additional booster vaccination during follow-up. In one patient, antibodies remained present after 24 weeks. However, the concentration of antibodies on this last study visit was nearly negative (35.8 BAUmL^-1^).

### Population PK analysis

Data from 42 patients were used for constructing of the population PK model. The Nab-titers obtained after dose administration were most adequately described using a two-compartment model with a mixed residual error model and IIV associated with CL, V1, and V2 (Table 2). The latter model was, subsequently, used for the covariate analysis. In the covariate analysis, lean body weight allowed to explain 6.5% of the estimated IIV for V1 using a power relationship most adequately. Moreover, a dichotomous covariate relationship distinguishing between the administration of ConvP or COVIg allowed explaining 15.3% of the IIV estimated for V2. Using the latter covariate relationship, the value for V2 was increased by a factor of 1.99 when ConvP was administered. This reduced the population PK parameter value of the base model estimated for V2 from 2700 mL to 1640 mL.

**Table 2.** Estimated population PK parameters for the structural model, final model, and bootstrap analysis.

|  | Base model |  |  | Final model |  |  | Bootstrap analysis |  |
| --- | --- | --- | --- | --- | --- | --- | --- | --- |
|  | Estimate | RSE (%) | Shr. (%) | Estimate | RSE (%) | Shr. (%) | Estimate | 95% CI |
| **Structural model** |  |  |  |  |  |  |  |  |
| Clearance (CL; mLh<sup>-1</sup>) | 9.14 | 6.7 |  | 9.1 | 6.6 |  | 9.2 | [8.0 - 10.4] |
| Volume of central compartment (V1; mL) | 3340 | 5 |  | 3790 | 4.9 |  | 3799 | [3428 - 4197] |
| Inter-compartmental clearance (Q; mLh<sup>-1</sup>) | 27.5 | 12.3 |  | 27.0 | 12.9 |  | 27.5 | [22.0 - 34.7] |
| Volume of peripheral compartment (V2; mL) | 2700 | 8.2 |  | 1640 | 15.4 |  | 1672 | [1198 - 2250] |
| **Inter-individual variability (%CV)** |  |  |  |  |  |  |  |  |
| IIV on CL | 46.1 | 8.6 | 0.1 | 44.3 | 8.5 | 0.4 | 45.8 | [37.3 – 54.5] |
| IIV on V1 | 33.8 | 11.1 | 0.9 | 27.3 | 13 | 2.3 | 26.9 | [20.2 – 34.1] |
| IIV on V2 | 44.7 | 15.6 | 11.5 | 29.2 | 19.6 | 17 | 29.6 | [17.5 – 43.0] |
| **Residual variability** |  |  |  |  |  |  |  |  |
| Additive residual error (BAUmL<sup>-1</sup>) | 3.82 | 12 |  | 3.75 | 10.9 |  | 3.75 | [2.685 - 4.515] |
| Proportional residual error (%CV) | 9.53 | 8 |  | 9.68 | 7.9 |  | 9.39 | [7.9 – 10.9] |
| **Covariate relationships** |  |  |  |  |  |  |  |  |
| V1 – Lean body weight | - |  |  | 0.538 | 26.4 |  | 0.546 | [0.260 - 0.860] |
| V2 – If using COVlg | - |  |  | 1.99 | 16.1 |  | 1.96 | [1.39 – 2.76] |
| **Model characteristics** |  |  |  |  |  |  |  |  |
| Objective function value | 2227.7 |  |  | 2194.0 |  |  | 2172.8 | [1993.0 - 2341.9] |
| Condition number (<1000) | 90.1 |  |  | 220.2 |  |  |  |  |

RSE indicates relative standard error; Shr., shrinkage; CI, confidence interval as obtained using the 2.5<sup>th</sup> and 97.5<sup>th</sup> percentiles from the non-parametric distributions; CV, coefficient of variation; BAU, binding antibody units.
in which the $\eta_i$ represents the random effect for an individual patient associated to a PK parameter. The covariate value CONVP associated to the PK parameter V2 was 1 in the case ConvP and 0 in the case COVlg was administered. The bootstrap analysis was conducted using 2000 replicated datasets.

In the group receiving high Nab-titer COVIg, anti-SARS-CoV-2 antibodies were measurable for a median duration of 41 (27 – 56) and 56 (52 – 84) days after 150 or 300 mL, respectively. Antibodies remained detectable for a longer time in patients who received ConvP than those who received COVIg independent of the administered volume and Nab-titer (Table 2). However, the median elimination half-life of ConvP and COVIg was comparable with 18.6 days and 20.3 days, respectively (Table S4). As expected, the peak Nab-titer was highest in the group that received 600mL ConvP with a Nab-titer of 500 IUmL^-1^. Duration of seropositivity for anti-SARS-CoV-2 antibodies and peak antibody concentration are presented in Table 2.

### Model evaluation

In Figure 2, the goodness-of-fit (GOF) plots for the final model are shown in which the adequacy of the model predictions is demonstrated. Both the individual and population predictions of the neutralizing antibody levels were symmetrically distributed around the line of identity (y=x), showing that accurate predictions of the Nab-titers from ConvP and COVIg were obtained using the final model. Moreover, the prediction-corrected visual predictive check (pcVPC) (Figure 3A) showed the accuracy of the final model as all quantiles from the measured Nab-titers (solid lines) were within their respective prediction intervals (shaded areas). However, when presented on the logarithmic scale the latest measured Nab-titers were slightly above the simulated Nab-titers (Figure 3B). The latter was due to the lowest amount of measured Nab-titers being present for that time period.

**Figure 2.**
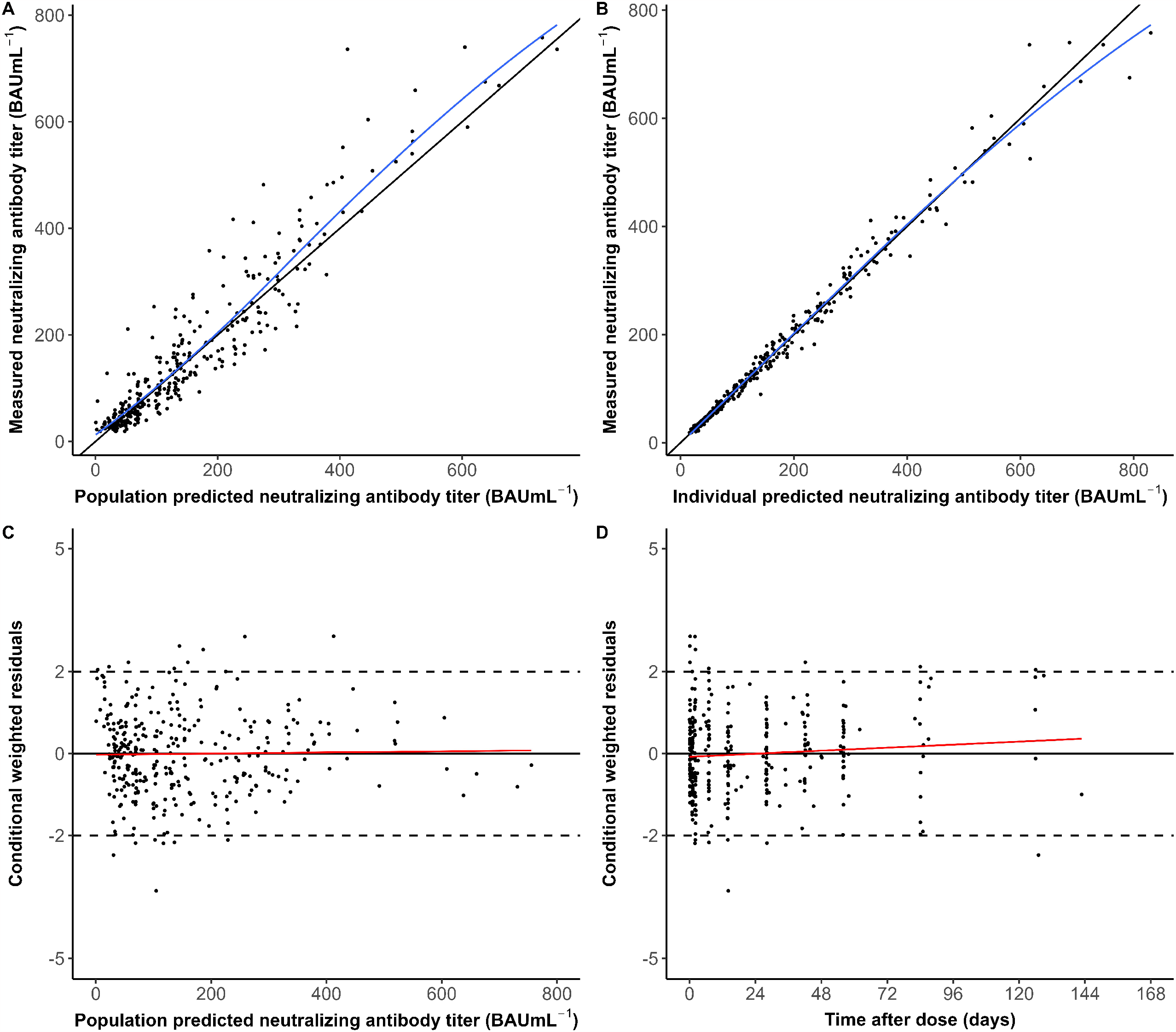
Goodness-of-fit plot from the final model. (A) Population predicted versus measured neutralizing antibody levels as quantified using binding antibody units per milliliter (BAUmL^-1^). (B) Individual predicted versus measured neutralizing antibody levels. (C) Conditional weighted residuals (CWRES) versus population predicted neutralizing antibody levels. (D) CWRES versus time after dose administration. In Figure A and B, the blue line depicts the locally weighted smoothing (LOESS) line, whereas in Figure C and D the red lines are the linear regression line.

**Figure 3.**
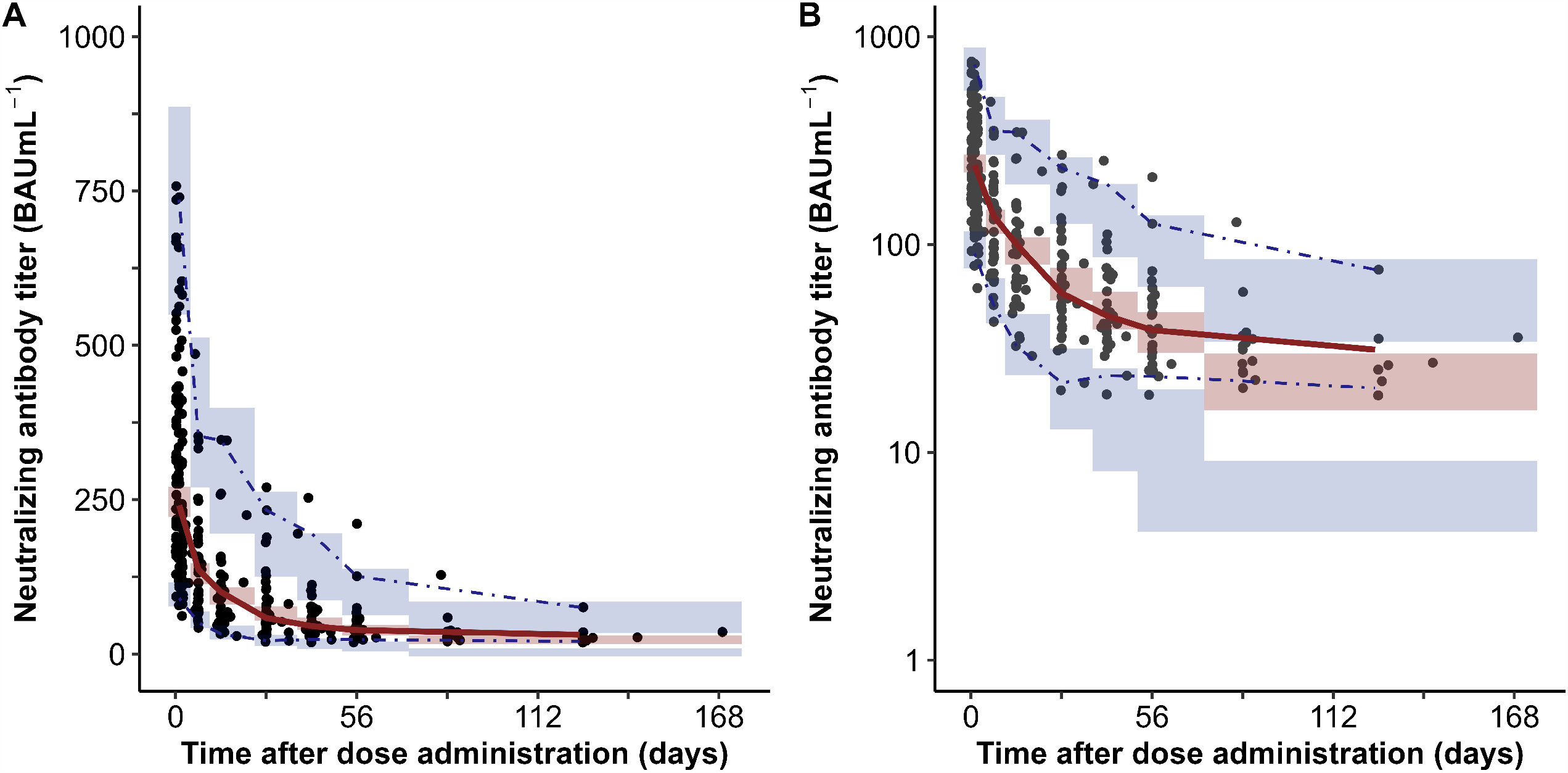
Prediction-corrected Visual Predictive Check from the final model. A) pcVPC with the measured and simulated neutralizing antibody (Nab) titers on a linear scale (B)) pcVPC with the observed and simulated neutralizing antibody (Nab) titers on a logarithmic scale. Black dots represent the measured neutralizing antibody levels for all patients. Solid grey line represents the median and the dashed grey lines represent the 2.5th and 97.5th quantiles for the measured neutralizing antibody levels. Red and blue-shaded areas show the 95% confidence intervals for the simulated neutralizing antibody titers of the individual patients, as obtained by 2000 Monte Carlo simulations using the final model.

The bootstrap analysis showed that the model parameters were adequately estimated, as all median parameter estimates from the bootstrap were similar to that of the final model.

### Dosing regimens simulations

Figure 4 depicts the Monte Carlo simulations evaluating the optimal dosing regimen of ConvP and COVIg. When dosing with 600mL of ConvP every 56 days (8 weeks), none of the simulated plasma Nab-titers achieved the 90% probability target attainment (PTA) for the 300 BAUmL^-1^ threshold. However, reducing the dosing interval to 28 days and using ConvP with a Nab-titer of at least 12,000 BAUmL^-1^ led to a 90% or higher PTA. For the lowest target of 100 BAUmL^-1^, the 90% PTA could be achieved with longer dosing intervals as long as ConvP or COVIg with an extremely high Nab-titer can be used (e.g. dosing every 8 weeks with 32,000 BAUmL^-1^ or more).

**Figure 4.**
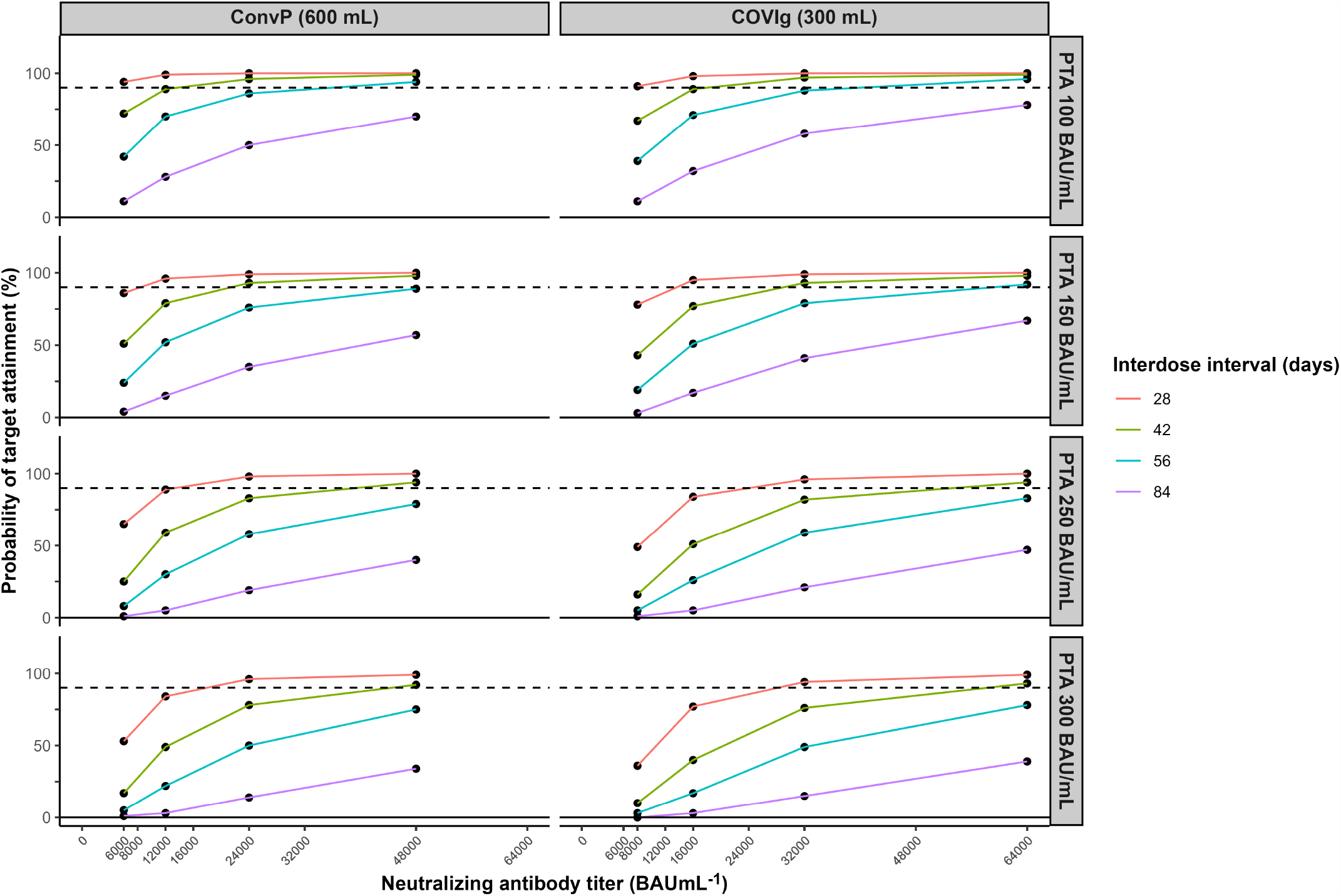
Probability of target attainment of simulated dosing regimens for ConvP and COVIg. For obtaining prediction for the neutralizing antibody levels, Monte Carlo simulations (n=2000) were applied using the final model. Each row of figures depicts the probability of the target attainment (PTA, %) for different target levels in BAUmL^-1^ in the serum of the recipient, as displayed in the facet header. The dashed line depicts 90% PTA, which was considered as the cut-off for having a protective effect. The volume of ConvP was set at 600 mL and for COVIg of 300 mL.

### Secondary endpoint

#### Breakthrough infections

Three patients had a breakthrough COVID-19 infection at a time when infused anti-spike antibodies were detectable. Two of them had received ConvP and one COVIg (Table 3). These breakthrough infections occurred with the most recently measured Nab-titer preceding the infection being 74.5 BAUmL^-1^, 51.2 BAUmL^-1^ and 68.6 BAUmL^-1^. These antibody titers were measured 31, 27, and two days before the first day of symptoms respectively. One of these patients required hospitalization. This patient was previously treated with anti-CD20 agents and was infected with the Delta (AY.9.2) variant. Treatment with casirivimab/imdevimab and dexamethasone was applied due to hypoxemia requiring supplemental oxygen. After five days, the patient was discharged. The two other patients were infected in the first trimester of 2022 at the time when Omicron BA.1 and BA.2 were the dominant strains in the Netherlands. These patients made a full recovery without hospital admission.

**Table 3.**
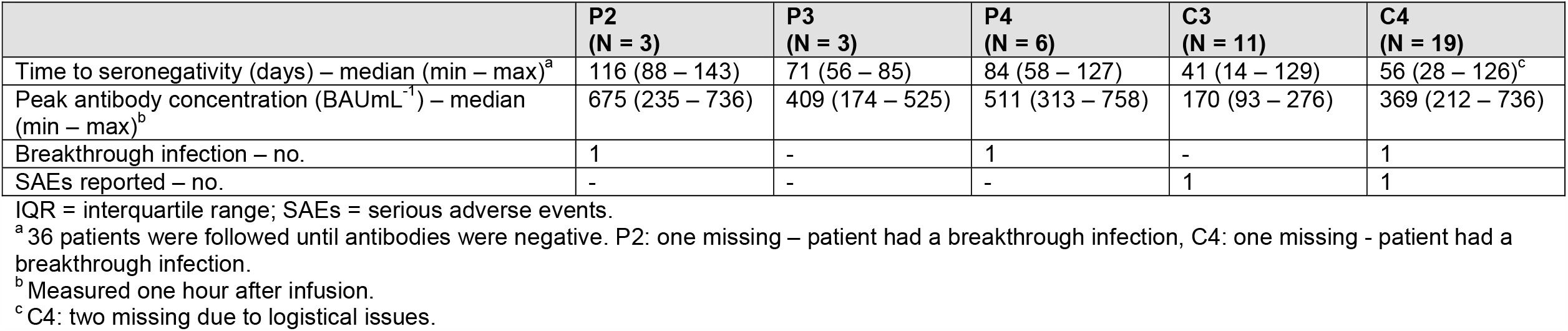
Outcome per subgroup.

|  | **P2**<br>**(N = 3)** | **P3**<br>**(N = 3)** | **P4**<br>**(N = 6)** | **C3**<br>**(N = 11)** | **C4**<br>**(N = 19)** |
| --- | --- | --- | --- | --- | --- |
| Time to seronegativity (days) – median (min – max)<sup>a</sup> | 116 (88 – 143) | 71 (56 – 85) | 84 (58 – 127) | 41 (14 – 129) | 56 (28 – 126)<sup>c</sup> |
| Peak antibody concentration (BAUmL<sup>-1</sup>) – median (min – max)<sup>b</sup> | 675 (235 – 736) | 409 (174 – 525) | 511 (313 – 758) | 170 (93 – 276) | 369 (212 – 736) |
| Breakthrough infection – no. | 1 | - | 1 | - | 1 |
| SAEs reported – no. | - | - | - | 1 | 1 |

IQR = interquartile range; SAEs = serious adverse events.
<sup>a</sup> 36 patients were followed until antibodies were negative. P2: one missing – patient had a breakthrough infection, C4: one missing - patient had a breakthrough infection.
<sup>b</sup> Measured one hour after infusion.
<sup>c</sup> C4: two missing due to logistical issues.

#### Serious adverse events

The administration of ConvP and COVIg was deemed safe. A SAE was reported in two patients (Table 3). However, these SAEs were unrelated to the studied products and were reported as an episode of hospitalization. The first patient was hospitalized due to COVID-19, and the second patient was due to a community-acquired pneumonia. Both patients recovered completely.

## Discussion

In this study, a population PK model predicting the Nab-titers after ConvP and COVIg administration was constructed. The adequacy of the model predictions was demonstrated using GOF plots and a pcVPC. Furthermore, a bootstrap analysis showed the robustness of the parameter estimates from the final model. Lean body weight was associated with V1. Concerning V2, its value increased approximately two times with administration of ConvP as compared with COVIg. Finally, Monte Carlo simulations showed that monthly dosing of ConvP with very high-titer ConvP (12,000 BAUmL^-1^) could attain the 90% PTA for the 300 BAUmL^-1^ target. To our knowledge, this is the first population PK model predicting Nab-titer after the administration of ConvP or COVIg.

In general, the elimination half-life of intravenous immunoglobulins ranges from 7 to 21 days. However, less is known about the elimination half-life of IgG subgroups targeting a specific antigen such as the spike protein of SARS-CoV-2.(28, 31) Before this trial, only a few studies have been performed evaluating the pharmacokinetic and pharmacodynamic characteristics of these pAbs. In a hamster model very high-titer COVIg (pseudovirus virus neutralization 50% titer: 1/1240) had a median elimination half-life of 124 hours.(32) In children, Gordon et al. investigated the PK of high-titer ConvP (Nab-titer of 1/320 anti-spike IgG, Euro-Immun) and found an elimination half-life for anti-SARS-CoV-2 IgG of 15 days whereas the median elimination half-life of pAbs in our population ranged between 18.6 and 20.6 days.(29)

A two-compartment model described the measured Nab-titers most adequately. In contrast to our model, Gordon et al. did not find body weight associated with clearance of the antibodies.(29) During the first elimination phase, a rapid decline in Nab-titers was observed in the first days after dose administration. During the distribution phase, IgG leaves the blood vasculature into lymph and extracellular compartments and slowly diffuses back into the blood circulation.(28) This rapid decline in Nab-titer may pose a problem for attaining higher Nab-titers for a long period and, thus, compromising the use in a prophylactic setting.(33) Adjustment of the Fc-receptor in antibodies is a strategy performed in mAbs and can prolong the elimination half-life of these antibodies.(31) Unfortunately, this is not possible in donor-based pAbs. Aside from this rapid decline, dose dilution in the systemic circulation is another factor that brings the need for very high-titer therapy.(33) In this study, the peak antibody measured 1 hour after administration in the blood of participants was 11 times lower than the titer in the ConvP or COVIg unit but with a broad IQR of 5 to 20 times. This was also observed by Shoham et al.(34) Although elimination half-life of pAbs is long, both the rapid decline in titer and dose dilution are factors that should be taken into account during the practical application of pAb-based prophylaxis in high-risk patients.

In this study, patients who were likely to or had been proven to lack an endogenous anti-SARS-CoV-2 antibody production as a result of B-cell depleting or B -and T-cell suppressive therapies were recruited. In this way, the efficacy of ConvP and COVIg in this patient group could be investigated. However, as the study had to be discontinued prematurely and only three patients had a breakthrough infection during follow-up, no definite conclusions about the titers needed for protection can be drawn. Furthermore, all 3 breakthrough infections occurred at a time when non-ancestral variants were circulating to which the study products had reduced activity.

Since the start of the pandemic, many clinical studies on the efficacy of ConvP and to a lesser extend also COVIg as a treatment for SARS-CoV-2 infected patients have been performed. The results of these studies were mostly disappointing because in hospitalized non-immunocompromised patients no clear beneficial effect was observed.(13, 16, 35-37) However, most of these trials were performed with plasma from convalescent and non-vaccinated donors. Therefore, donors with extremely high Nab-titers were rare. We previously summarized the available evidence on optimal dosing of ConvP and concluded that patients were underdosed in almost all of these trials.(19) Furthermore, it has become clear that antibody-based therapy works best when given in the first days after symptom onset. Indeed, in a recent meta-analysis of the 6 double-blind randomized trials on ConvP performed in outpatients with <8 days of symptoms, a significant reduction in hospital admission was only observed when the intervention was given in the first 5 days of symptoms and when plasma with the highest antibody titers was used.(17)

To explore the clinical application of the prophylactic use of ConvP and COVIg, a PTA was estimated with a Monte Carlo simulation using 4 antibody titer targets ranging from 100 to 300 BAUmL^-1^. These titers were deemed relevant, as Feng et al. showed that a titer of 264 BAUmL^-1^ was associated with 80% vaccine efficacy whereas Goldblatt et al. and Dimeglio et al. reported 150 BAUmL^-1^ as sufficient for offering protection.(38-40) However, protection against infection does not only come from humoral immunity which may implicate that in patients with a B and T-cell deficiency higher titers may be required. Also, subsequent variants of concern are often much more resilient to vaccine-induced antibodies and higher titers are necessary to offer protection.(41) Dimeglio et al. showed that achieving titers over 20.000 BAUmL^-1^ are necessary to achieve at least 80% of protection against Omicron infections.(42) In the current simulation, a PTA of 300 BAUmL^-1^ was achieved by administering ConvP or COVIg with at least 12,000 BAUmL^-1^ every 4 weeks.

This study has its limitations. In SARS-CoV-2 uninfected patients, the final model can be used to predict Nab-titers over time after an infusion of ConvP or COVIg. However, in patients infected with COVID-19, exogenous antibodies probably have a shorter half-life due to the direct antibody-antigen binding.(32) Also, with every new variant that occurs, the correlation of a SARS-CoV2 spike antibody titer (in BAUmL^-1^) with in vitro neutralization of this new variant should be evaluated again. This means that for new variants, much higher targets (e.g. 10.000 rather than 300 BAUmL^-1^) may be necessary to result in any relevant protection. Fortunately, many plasma donors now have acquired immunity from a combination of infection, vaccination and booster vaccination. Therefore, donors with extremely high Nab-titers are readily identifiable. Unfortunately, due to the rapidly evolving variant landscape of SARS-CoV-2 and the vaccination uptake, the study was discontinued prematurely and only three breakthrough infections were detected. Therefore, a protective titer could not be estimated.

In conclusion, this is the first dose-finding study in which a population PK model describing Nab-titers after ConvP and COVIg administration was constructed. Lean body weight and the type of pAbs allowed to explain a part of the IIV for V1 and V2, respectively. This population PK model may be a valuable tool for designing trials during future viral pandemics at the time when application of ConvP or COVIg is considered as a prophylactic or therapeutic intervention.

## Study highlights

### What is the current knowledge on the topic?

Although the pharmacokinetics of immunoglobulins have been elucidated, less is known about the pharmacokinetics of target-specific polyclonal antibodies.

### What question did this study address?

As the most optimal dosing regimens for convalescent plasma and hyperimmune globulins containing anti-SARS-CoV-2 antibodies are unknown, a population pharmacokinetic analysis was performed.

### What does this study add to our knowledge?

In this study, the population pharmacokinetic model of convalescent plasma and hyperimmune globulins containing anti-SARS-CoV-2 antibodies is established for the first time. This way we were able to show that very high-titer agents are needed to achieve an optimal dosing regimen for these products.

### How might this change clinical pharmacology or translational science?

The population PK model can be applied for designing trials during future pandemics if application of polyclonal antibody therapy is considered as prevention against or treatment of viral infections.

## Supporting information

Supplemental figure 1: Trial design

Supplementary methods S2: Neutralizing antibody measurement

Supplementary table S3: Baseline characteristics per subgroup

Supplementary table S4. Terminal elimination half-life and AUC for ConvP and COVIg

## Data Availability

All data produced in the present study are available upon reasonable request to the authors

## Acknowledgements

We thank all volunteers that participated in the trial as well as all plasma donors.

## Author contributions

BR, BK, SH, and TP designed the research. Research was performed by SH and supervised by BR. SH and TP analyzed the data and TP constructed the population PK model. The manuscript was written by SH and TP, the manuscript was reviewed by BR, BK, FS, IKB, and CG. BR did the overall supervision of the project.

## Supplementary information titles

**Supplementary figure S1: Trial design**

**Supplementary methods S2: Neutralizing antibody measurement**

**Supplementary table S3: Baseline characteristics per subgroup**

**Supplementary table S4. Terminal elimination half-life and AUC for ConvP and COVIg**.

