## Supplementary figures and images for "Dosing of convalescent plasma and hyperimmune anti-SARS-CoV-2 immunoglobulins: a phase I/II dose finding study"

### Supplemental figure 1: Trial design

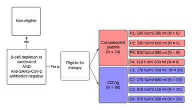
