## Supplementary methods S2: Neutralizing antibody measurement for "Dosing of convalescent plasma and hyperimmune anti-SARS-CoV-2 immunoglobulins: a phase I/II dose finding study"

A serum sample from each plasma donor taken on the day of plasma donation was sent to the National Institute for Public Health and the Environment (Dutch: RIVM) lab for virus neutralization testing. Duplicates of two-fold serial dilutions (starting at 1:10) of heat-inactivated sera (30 m, 56°C) were incubated with 100 median tissue culture infectious dose of SARS-CoV-2 strains hCoV-19/Netherlands/ZuidHolland_10004/2020, D614G (WT) and hCoV19/Netherlands/ NoordHolland_10159/2021 (B.1.351, EVAg, catalog no. 014 V-04058) at 35°C for 1 hour in 96-well plates. Vero E6 cells were added in a concentration of 20,000 cells per well and were incubated for 72 hours at 35°C. The serum virus neutralization titer was defined as the reciprocal value of the sample dilution that showed a 50% protection of virus growth. Samples with titers of ≥20 were defined as SARS-CoV-2 seropositive. To facilitate conversion to International Units (IU), a calibrated panel of plasma samples containing a dilution series of a high titer convalescent plasma calibrated in IUmL^-1^ using the standard 20/130 obtained from the National Institute for Biological Standards and Control (NIBSC, United Kingdom). 2 Experimental neutralization titers were converted to IUmL^-1^ using the following regression formula (IUmL^-1^ = 4160/(2^(Log2 (experimentalID50-11.832)/-1.146) derived from assay calibration with the pre-quantified control.
