## Supplementary table S3: Baseline characteristics per subgroup for "Dosing of convalescent plasma and hyperimmune anti-SARS-CoV-2 immunoglobulins: a phase I/II dose finding study"

**Supplementary table S3: baseline characteristics subgroups**

| **Baseline characteristics** | **ConvP 500 IUmL^-1^ 600 mL**  **n = 3** | **ConvP 910 IUmL^-1^ 300 mL**  **n = 3** | **ConvP 910 IUmL^-1^ 600 mL**  **n = 6** | **COVIg 910 IUmL^-1^ 150 mL**  **n = 11** | **COVIg 910 IUmL^-1^ 300 mL**  **n = 19** |
| --- | --- | --- | --- | --- | --- |
| Age (yr) – median (min - max) | 61 (35 – 63) | 57 (41 – 64) | 60 (43 – 72) | 62 (31 – 70) | 61 (41 – 74) |
| Male gender – no. (%) | - | 2 (66.7) | 3 (50.0) | 6 (54.5) | 11 (57.9) |
| Race   - Caucasian - African - Asian | 3 (100.0)  -  - | 2 (66.7)  1 (33.3)  - | 5 (83.3)  -  1 (16.7) | 11 (100.0)  -  - | 16 (842)  1 (5.3)  1 (5.3) |
| BMI – median (min - max) | 30 (18 – 34) | 28 (23 – 43) | 24 ( 21 – 33) | 24 (16 – 34) | 28 (22 – 43) |
| Body weight – median (min - max) | 81 (57 – 88) | 82 (62 – 145) | 80 (67 – 108) | 86 (48 – 99) | 83 (62 – 144) |
| Body length – median (min - max) | 165 (160 – 177) | 172 (164 – 183) | 180 (158 – 189) | 178 (167 – 191) | 177 (156 – 186) |
| Cause for immunocompromised state – no. (%)   - Rituximab - SOTx - Ocrelizumab - HSCT - CVID - Other reason | 2 (66.7)  -  -  -  1 (33.3)  - | 2 (66.7)  -  1 (33.3)  -  -  - | 4 (66.7)  1 (16.7)  1 (16.7)  -  -  - | 7 (63.6)  1 (9.1)  1 (9.1)  2 (18.2)  -  - | 11 (57.9)  5 (26.3)  1 (5.3)  2 (10.5) |
| Vaccination status   - Full vaccination ^a^ - Third vaccination ^b^ | 2 (66.7)  2 (100.0) | 3 (100.0)  1 (33.3) | 5 (83.3)  1 (20.0) | 9 (81.8)  3 (33.3) | 19 (100)  1 (5.3) |
| Laboratory findings – median (min – max)   - HCT - WBC - Lymphocytes - B-cells - T-cells - CD4+ - CD8+ - NK cells | 37 (36 – 43)  5300 (4400 – 12000)  1250 (960 – 1580)  0 (0 – 0)  1225 (1170 – 1280)  735 (710 – 760)  490 (470 – 510)  265 (80 – 450) | 47 (38 – 49)  4400 (3800 – 4500)  1020 (590 – 1070)  0 (0 – 0)  730 (580 – 740)  490 (100 – 540)  220 (200 – 480)  240 (110 – 360) | 42 ( 37 – 45)  4550 (3100 – 8600)  955 (530 – 1230)  0 (0 – 70)  715 (300 – 1240)  505 (160 – 1080)  185 (120 – 370)  260 (40 – 290) | 43 (25 – 45)  5500 (2500 – 14500)  880 (160 – 2800)  0 (0 – 40)  720 (80 – 2290)  440 (40 – 1610)  230 (40 – 700)  150 (50 – 420) | 42 (36 – 48)  6500 (3600 – 18100)  1100 (720 – 9880)  0 (0 – 6430)  830 (460 – 1600)  570 (120 – 1130)  350 (90 – 720)  190 (60 – 470) |

ConvP = convalescent plasma; COVIg = hyperimmune anti-SARS-Cov-2 globulins; CVID = common variable immune deficiency; HSCT = hematopoietic stem cell transplant; NK = natural killer; SOTx = solid organ transplant.

a : Two mRNA vaccines, two vaccines by AstraZeneca or one Johnson & Johnson.

b : Second vaccination (mRNA) in case of vaccination with Johnson & Johnson.
