## Supplementary table S4. Terminal elimination half-life and AUC for ConvP and COVIg for "Dosing of convalescent plasma and hyperimmune anti-SARS-CoV-2 immunoglobulins: a phase I/II dose finding study"

|  | **ConvP** | | **COVIg** | |
| --- | --- | --- | --- | --- |
|  | ***t*_1/2_ (days)** | **AUC (Nab-titer * days / mL)** | ***t*_1/2_ (days)** | **AUC (Nab-titer * days / mL)** |
| **Minimum** | 9.31 | 1859 | 12.7 | 909 |
| **25% quartile** | 16.5 | 5056 | 17.4 | 2004 |
| **Median** | 18.6 | 6579 | 20.3 | 3458 |
| **Mean** | 21.1 | 9409 | 22.3 | 3508 |
| **75% quartile** | 22 | 10792 | 25.5 | 4353 |
| **Maximum** | 46.2 | 28881 | 50.6 | 8055 |

* Nab-titer was measured in BAUmL^-1^.
